# COVID-19 infection wave mortality from surveillance data in the Philippines using machine learning

**DOI:** 10.1101/2023.11.28.23299037

**Authors:** Julius R Migriño, Ani Regina U Batangan, Rizal Michael R Abello

## Abstract

**Objective:** The Philippines has had several COVID-19 infection waves brought about by different strains and variants of SARS-CoV-2. This study aimed to describe COVID-19 outcomes by infection waves using machine learning.

**Methods:** We used a cross-sectional surveillance data review design using the DOH COVID DataDrop data set as of September 24, 2022. We divided the data set into infection wave data sets based on the predominant COVID-19 variant(s) of concern during the identified time intervals: ancestral strain (A0), Alpha/Beta variant (AB), Delta variant (D), and Omicron variant (O). Descriptive statistics and machine learning models were generated from each infection wave data set.

**Results:** Our final data set consisted of 3 896 206 cases and ten attributes including one label attribute. Overall, 98.39% of cases recovered while 1.61% died. The Delta wave reported the most deaths (43.52%), while the Omicron wave reported the least (10.36%). The highest CFR was observed during the ancestral wave (2.49%), while the lowest was seen during the Omicron wave (0.61%). Higher age groups generally had higher CFRs across all infection waves. The A0, AB and D models had up to four levels with two or three splits for each node. The O model had eight levels, with up to 16 splits in some nodes. Of the ten attributes, only age was included in all the decision tree models, while region of residence was included in the O model. F-score and specificity were highest using naïve Bayes in all four data sets. Area under the curve (AUC) was highest in the naïve Bayes models for the A0, AB and D models, while sensitivity was highest in the decision tree models for the A0, AB and O models.

**Discussion:** The ancestral, Alpha/Beta and Delta variants seem to have similar transmission and mortality profiles. The Omicron variant caused lesser deaths despite being more transmissible. Age remained a significant predictor of death regardless of infection wave. We recommend constant timely analysis of available data especially during public health events and emergencies.

## INTRODUCTION

The Philippines has been considered a hotspot for the coronavirus disease 2019 (COVID-19) in the Western Pacific region.^1^ As of December 1, 2022, the country’s Department of Health (DOH) has reported a total of 4,037,547 cases, including 64,658 reported deaths.^2^ Meanwhile, the World Health Organization (WHO) has tallied 639,572,819 confirmed cases and 6,615,258 deaths globally.^3^ The severe acute respiratory syndrome coronavirus 2 (SARS-CoV-2) is the primary etiologic agent of COVID-19 infection. The infection causes symptoms like cough, colds, fever, dyspnea and dysgeusia, and may progress to be more life-threatening if it presents with complications such as shock and organ failure. COVID-19 mortality is influenced by several factors like advanced age, sex, presence of pre-existing comorbid illness, and history of smoking and alcohol consumption.^1^ More recently, studies have surfaced highlighting the differences in mortality rates among cases with different vaccination statuses^4,5^ and among those who were previously infected.^5^ According to SeyedAlighani and his colleagues (2021), mortality rates are additionally influenced by adequacy of health care delivery, political decisions, and epidemiological characteristics of the affected population.^6^

Generally, viruses evolve to become more transmissible, regardless of severity.^7^ The ancestral strain was the original SARS-CoV-2 virus which originated in China. The virus has been persistent in its infection rates due to its intrinsic capability to replicate and mutate. These spontaneous mutations are products of viral RNA replication errors within the host cell resulting in the appearance of multiple variants.^8^ As of December 2022, there have been five recognized circulating SARS-CoV-2 variants of concern (VOCs): Alpha, Beta, Gamma, Delta and Omicron. These VOCs have appeared in infection waves among different countries in varying timelines, but their designation as VOCs were December 2020 (Alpha and Beta), January 2021 (Gamma), May 2021 (Delta) and November 2021 (Omicron).^9^ Recent studies have characterized the different VOCs in terms of their transmissibility and severity. For instance, while the Delta variant evolved to become more transmissible than previous variants, several studies report similar hospitalization and mortality rates among the different infection waves.^10–12^ The Omicron variant, on the other hand, proved to be even more highly transmissible compared to the previous variants, but has shown the lowest hospitalization and mortality rates.^13^ The observed differences in transmission and severity among COVID-19 variants is possibly related to the increased immunity among the people infected, either through vaccination or previous infection waves.^7^

As of October 8, 2022, there had been a total of 22,400 SARS-CoV-2 sequences shared by the Philippines in the Global Initiative on Sharing All Influenza Data (GISAID) COVID-19 sequence repository, which accounts for 0.57% of all cases.^14^ Tracking of relative frequencies of variants from sequenced COVID-19 cases show estimated time frames of the upsurge of specific variants: the ancestral strain was predominant (i.e., made up more than 50% of all sequenced samples) until about February 2021; the Alpha and Beta variants were concurrently predominant starting March until June 2021; the Delta variant was predominant from July until November 2021; starting from December 2021 until present, the Omicron variant and its subvariants were predominant.^15^

Machine learning is often used for health in the analysis of large datasets and the prediction of outcomes based on a variety of inputs. Such applications of machine learning include identification of disease from clinical symptoms or laboratory results, as well as in treatment of diseases and facilitation of administrative processes. Such techniques have been used to aid in treatment of cancer, pneumonia, diabetes and other diseases, including COVID-19, wherein they can give more than 90% accuracy in prediction and forecasting.^16^

Early prediction of COVID-19 mortality risks may help mitigate the effect of the pandemic by providing evidence for efficient resource allocation and proper patient treatment plans,^17^ and has been the topic of several researches.^17–19^ Most studies relied on medical records from admitted patients, relying on demographic, clinical and laboratory features to generate predictive models for patient prognosis. Some examples of machine learning algorithms used in COVID-19 research include logistic regression,^18^ support vector machines,^17^ and decision tree ensembles (e.g., CatBoost, XGBoost, Random Forest).^19^ A previous study utilized a publicly available national surveillance dataset to predict COVID-19 mortality in the Philippines and identified age and history of hospital admission as significant predictors of disease outcome, but the study was limited to the early part of the pandemic wherein only the ancestral strain was present in the population.^20^ This study aimed to describe COVID-19 outcomes by infection waves using machine learning.

## METHODS

The study utilized a cross-sectional, documents review design. Data from the publicly available DOH COVID Data Drop database for September 24, 2022^2^ was utilized in this study. The database represented all reported COVID-19 cases by reverse transcription polymerase chain reaction of respiratory swabs and was updated daily by the DOH Epidemiology Bureau. The raw data set contained 3 934 777 cases and 22 attributes. Exploratory analysis was performed to screen cases and attributes in the raw data set. Ten attributes were included in the model generation which included *Age, Sex, Admitted, RegionRes, ProvRes, CityMunRes, BarangayRes, Quarantined, Pregnanttab* and *RemovalType*. Another attribute *Age_Group* was generated to reclassify *Age* into nine bins based on the US CDC classification for descriptive statistics. Another attribute, *DateRepConf*, was retained only for splitting of the data sets (below). Missing values for *Pregnanttab* were recoded as “(N/A)” for cases with *Sex*=MALE. Missing values for *BarangayRes, CityMunRes* and *ProvRes* were recoded as “ROF” for all cases where *RegionRes*=“ROF”. Cases with missing values for *Age* and *RemovalType* were dropped from the data set to generate the final data sets. Details of the exploratory analysis can be found in **Supplementary Information A**.

The final dataset was split into four (4) data sets according to *DateRepConf*, where each data set represents the predominant COVID-19 variant(s) of concern during those time intervals as reported by GISAID^15^. The details of the four data sets are listed below:

1. A0 data set (predominant strain: ancestral; start date: January 30, 2020; end date: February 28, 2021);
2. AB data set (predominant variants: Alpha and Beta; start date: March 1, 2021; end date: June 30, 2021);
3. D data set (predominant variant: Delta; start date: July 1, 2021; end date: November 30, 2021);
4. O data set (predominant variant: Omicron; start date: December 1, 2021; end date: September 24, 2022)

Descriptive statistics such as means, standard deviations, frequencies, case fatality rates (CFR), t tests and Pearson’s χ^2^ tests were generated with StataCorp 2013 (Stata Statistical Software, Release 13; College Station, TX).

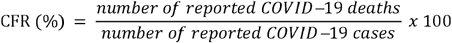

Attribute selection, random undersampling, hyperparameter optimizations, model generation, cross-validation and calculations of model performances were done in RapidMiner Studio 9.10.008 (rev: 68db53, platform: WIN64) (see **Supplementary Information B**). The attribute *RemovalType* was used as the outcome in all data sets. Attribute selection was done individually for all data sets using feature weights operators *weightbyGiniIndex* and *weightbyInformationGainRatio* to determine appropriate attributes to be included in the model generation. Grid optimizations of the hyperparameters for the decision tree operator *decisionTree* for all data sets were done by running fivefold cross-validation using the subprocess *optimizeParameters(Grid)* and operator *crossValidation*. Model generation was done for all four data sets using fivefold cross-validation using the optimized hyperparameters and *RemovalType*=DIED as the positive class set. Random undersampling (RUS) was done only on the training data sets for each fold, and was done using the *sample* operator to a) select all cases with *RemovalType*=DIED, and b) randomly select cases with *RemovalType*=RECOVERED using simple random sampling to achieve a 1:1 RECOVERED:DIED ratio. This training dataset was used to generate the decision tree models per fold. All cases in the testing data sets were used to validate each model.

The decision tree models generated per data set were extracted. Performance metrics such as area under the curve (AUC), accuracy, F-score, sensitivity and specificity were extracted from the cross-validation. Similar cross-validation operators were used to generate naïve Bayes and random forest models and performance metrics of all data sets. Receiver operating characteristic (ROC) curves for the three models were also generated using RapidMiner Studio 9.10.008 (rev: 68db53, platform: WIN64).

## RESULTS

### Description of cases

The final data set consisted of 3 896 206 cases (99.02% of all total reported cases from the raw data set) and 10 attributes including one label attribute (*RemovalType*). The A0, AB, D and O data sets comprised 14.68%, 21.45%, 36.31% and 27.56% of the reported cases in the final data set, respectively. The daily reported cases as well as the segmentation according to variants are visualized in **Fig. 1**. Of all reported cases, 98.39% recovered while 1.61% died. Among all reported deaths, the D data set contributed the most cases (43.52%) while the O data set contributed the least (10.36%). Among the four data sets, the highest CFR occurred during the first wave (2.49%) and the lowest during the Omicron wave (0.61%).the Alpha/Beta waves, reported cases were predominantly males, but the CFRs among males were higher than females across all four data sets. Cases with age over 85 years had the highest CFR among different age groups, while cases in the 5-17, 18-29 and 30-39 age groups had the lowest CFRs. Age-stratified CFRs in the Alpha/Beta, Delta and Omicron waves were lower compared to the ancestral wave across age groups (**Table 1**).

**Table 1.**
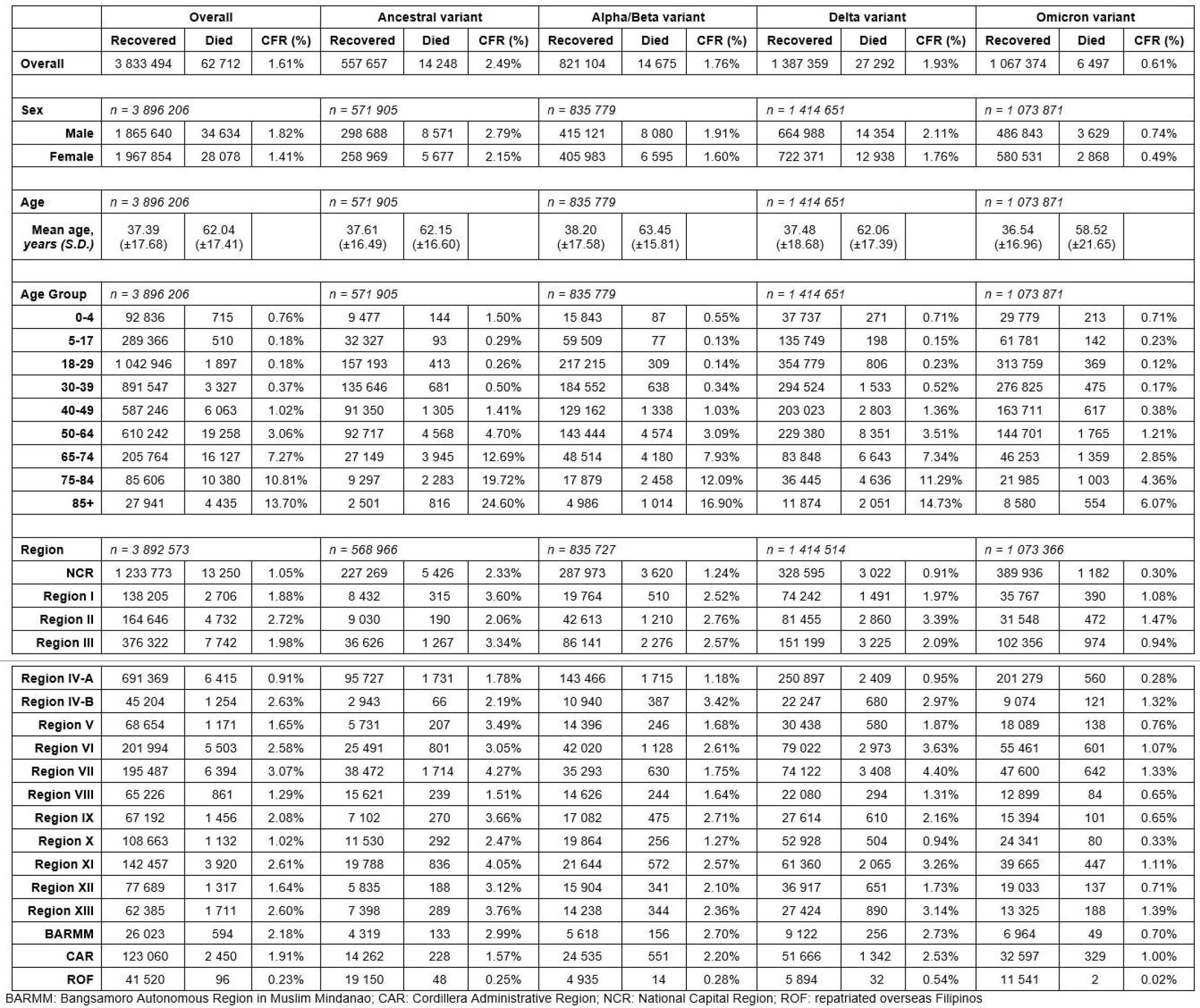
Demographic characteristics of reported cases (recovered or died) from the Philippines COVID Data Drop from September 24, 2022.

**Figure 1.**
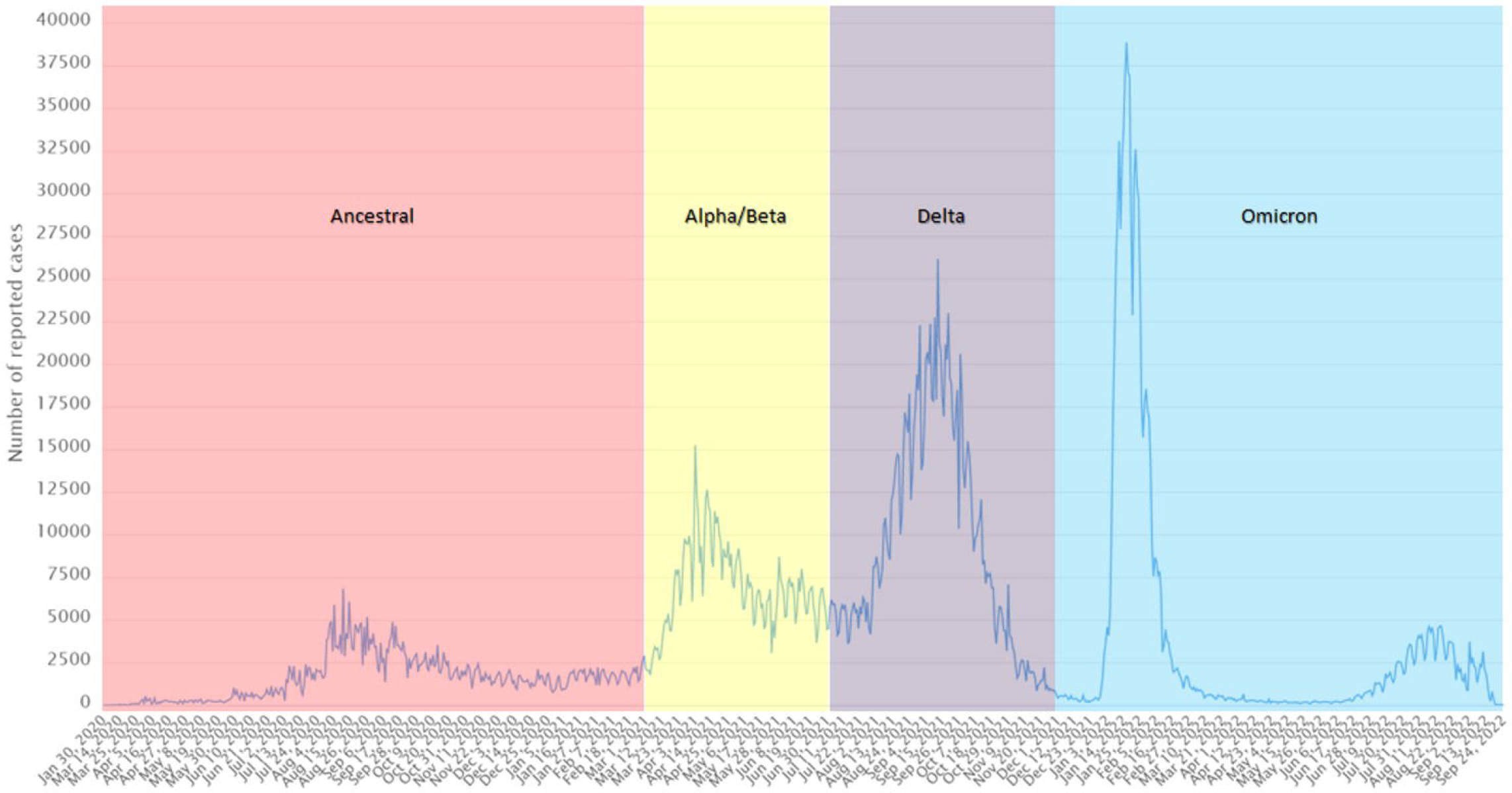
Reported COVID-19 cases in the Philippines by predominant variant

Based on disaggregation by region, the National Capital Region (NCR), Cordillera Autonomous Region (CAR), Region II and Region IV-A reported the highest case rates overall (9 304, 7 007, 4 603 and 4 323 cases per 100 000, respectively) and among most of the four data sets. The highest CFR was recorded in Region VII during the Delta wave (4.40%), while the lowest CFR was recorded in repatriated overseas Filipinos (ROF) during the Omicron wave (0.02%) (**Table 1**).

### Outcomes from machine learning models

Out of the nine non-outcome attributes retained for model generation, only *Age* and *Admitted* were included in the models for data sets A0, AB and D. For the data set O, *Age, Admitted* and *RegionRes* were included in the model. The models were trained and cross-validated with optimized hyperparameters detailed in **Supplementary Information B.3-5**.

In terms of performance, accuracy, F-score and specificity were highest using naïve Bayes in all four data sets. AUC was highest in the naïve Bayes models for the A0, AB and D data sets, while sensitivity was highest in the decision tree models for the A0, AB and O data sets (**Table 2**). The ROC curves for the naïve Bayes and random forest models were better compared to the ROC curve of the decision tree model (**Fig. 2**).

**Table 2.** Performance metrics for the three machine learning models: decision tree, naïve Bayes and random forest using the four modelling data sets and optimized hyperparameters.

| A0 dataset |  |  |  |  |  |  |  |  |  |  |
| --- | --- | --- | --- | --- | --- | --- | --- | --- | --- | --- |
| Model | AUC |  | Accuracy |  | F-score |  | Sensitivity |  | Specificity |  |
| Decision Tree | 0.789 | ± 0.004 | 74.06% | ± 0.79% | 13.88% | ± 0.35% | 83.86%<sup>a</sup> | ± 0.35% | 73.81% | ± 0.81% |
| Naïve Bayes | 0.877<sup>a</sup> | ± 0.004 | 80.25%<sup>a</sup> | ± 0.60% | 17.00%<sup>a</sup> | ± 0.36% | 81.16% | ± 0.55% | 80.23%<sup>a</sup> | ± 0.62% |
| Random Forest | 0.824 | ± 0.018 | 74.66% | ± 0.62% | 14.10% | ± 0.28% | 83.43% | ± 0.54% | 74.43% | ± 0.65% |
| AB dataset |  |  |  |  |  |  |  |  |  |  |
| Model | AUC |  | Accuracy |  | F-score |  | Sensitivity |  | Specificity |  |
| Decision Tree | 0.781 | ± 0.004 | 68.36% | ± 1.80% | 8.91% | ± 0.35% | 88.03%<sup>a</sup> | ± 1.21% | 68.00% | ± 1.85% |
| Naïve Bayes | 0.869<sup>a</sup> | ± 0.004 | 77.07%<sup>a</sup> | ± 0.08% | 11.22%<sup>a</sup> | ± 0.11% | 82.51% | ± 0.62% | 76.97%<sup>a</sup> | ± 0.07% |
| Random Forest | 0.798 | ± 0.003 | 68.76% | ± 1.11% | 8.99% | ± 0.22% | 87.79% | ± 0.90% | 68.42% | ± 1.14% |
| D dataset |  |  |  |  |  |  |  |  |  |  |
| Model | AUC |  | Accuracy |  | F-score |  | Sensitivity |  | Specificity |  |
| Decision Tree | 0.769 | ± 0.006 | 66.15% | ± 2.73% | 9.12% | ± 0.48% | 87.69% | ± 1.77% | 65.73% | ± 2.82% |
| Naïve Bayes | 0.844<sup>a</sup> | ± 0.003 | 74.74%<sup>a</sup> | ± 0.05% | 10.98%<sup>a</sup> | ± 0.08% | 80.73% | ± 0.58% | 74.62%<sup>a</sup> | ± 0.05% |
| Random Forest | 0.779 | ± 0.005 | 65.32% | ± 2.62% | 8.96% | ± 0.46% | 88.18%<sup>a</sup> | ± 1.73% | 64.87% | ± 2.70% |
| O dataset |  |  |  |  |  |  |  |  |  |  |
| Model | AUC |  | Accuracy |  | F-score |  | Sensitivity |  | Specificity |  |
| Decision Tree | 0.814 | ± 0.014 | 77.27% | ± 2.34% | 3.93% | ± 0.26% | 76.42%<sup>a</sup> | ± 2.82% | 77.28% | ± 2.37% |
| Naïve Bayes | 0.843 | ± 0.006 | 80.30%<sup>a</sup> | ± 0.24% | 4.38%<sup>a</sup> | ± 0.08% | 74.53% | ± 1.55% | 80.33%<sup>a</sup> | ± 0.24% |
| Random Forest | 0.844<sup>a</sup> | ± 0.006 | 78.32% | ± 1.02% | 4.09% | ± 0.13% | 76.25% | ± 1.61% | 78.33% | ± 1.04% |

<sup>a</sup> Highest values for each metric across all models

**Figure 2.**
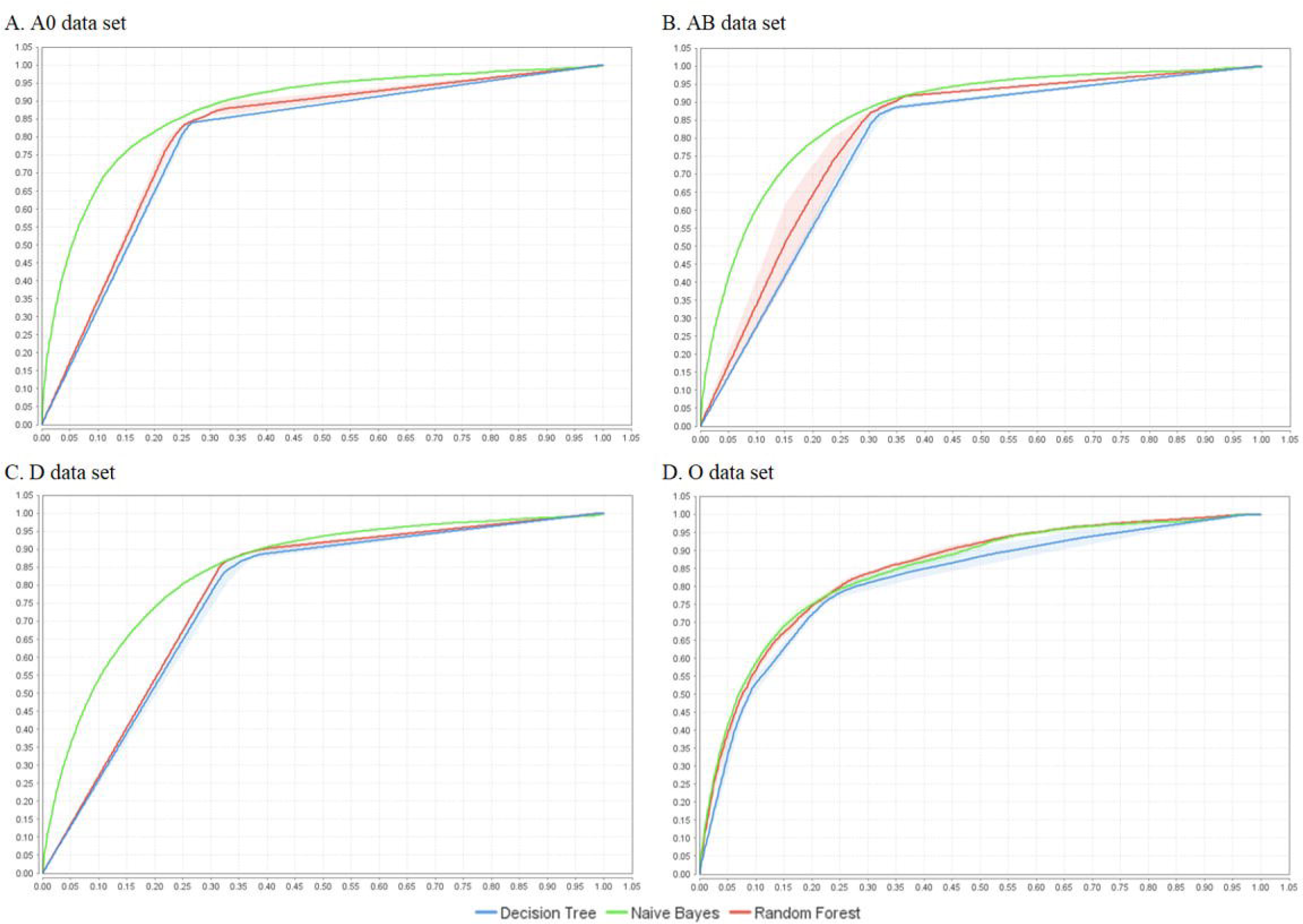
Receiver operating characteristic (ROC) curves for the machine learning models by data set: decision tree, naïve Bayes, random foresta^a^ ^a^The ROC curve plots a model’s sensitivity, or true positive rate, versus its false positive rate (one minus the specificity or true negative rate) as its discrimination threshold is varied. Generally, the closer the ROC curve is to the top left corner of the graph, the better the model.

The decision tree models for the A0 and AB data sets were similar: they were composed of three levels, with each node splitting into two branches (**Fig. 3A** and **Fig. 3B**, respectively). The D data set had four levels and had either two or three splits (**Fig. 3C**). The root node for the A0, AB and D datasets was *Age*, with the lowest split criterion in the D data set (41.5 years) and the highest in the A0 data set (47.5 years). Another split according to *Age* was also observed in all three data sets at *Age* = 0.5 years. The attribute *Admitted* also split the D data set for cases with *Age* <= 41.5 years (**Fig. 3C**). Majority of cases above the root node cutoffs died in all three data sets (A0 = 76.60%, AB = 72.93%, D = 70.21%), while majority of cases within or below the root node cutoff and above *Age* = 0.5 years recovered (A0 = 82.88%, AB = 86.19%, D = 86.39%). In the D data set, 64.04% of cases who had a history of hospital admission died. In the A0, AB and D data sets, majority of cases below *Age* = 0.5 years died (A0 = 65.88%, AB = 59.70%, D = 55.61%).

**Figure 3.**
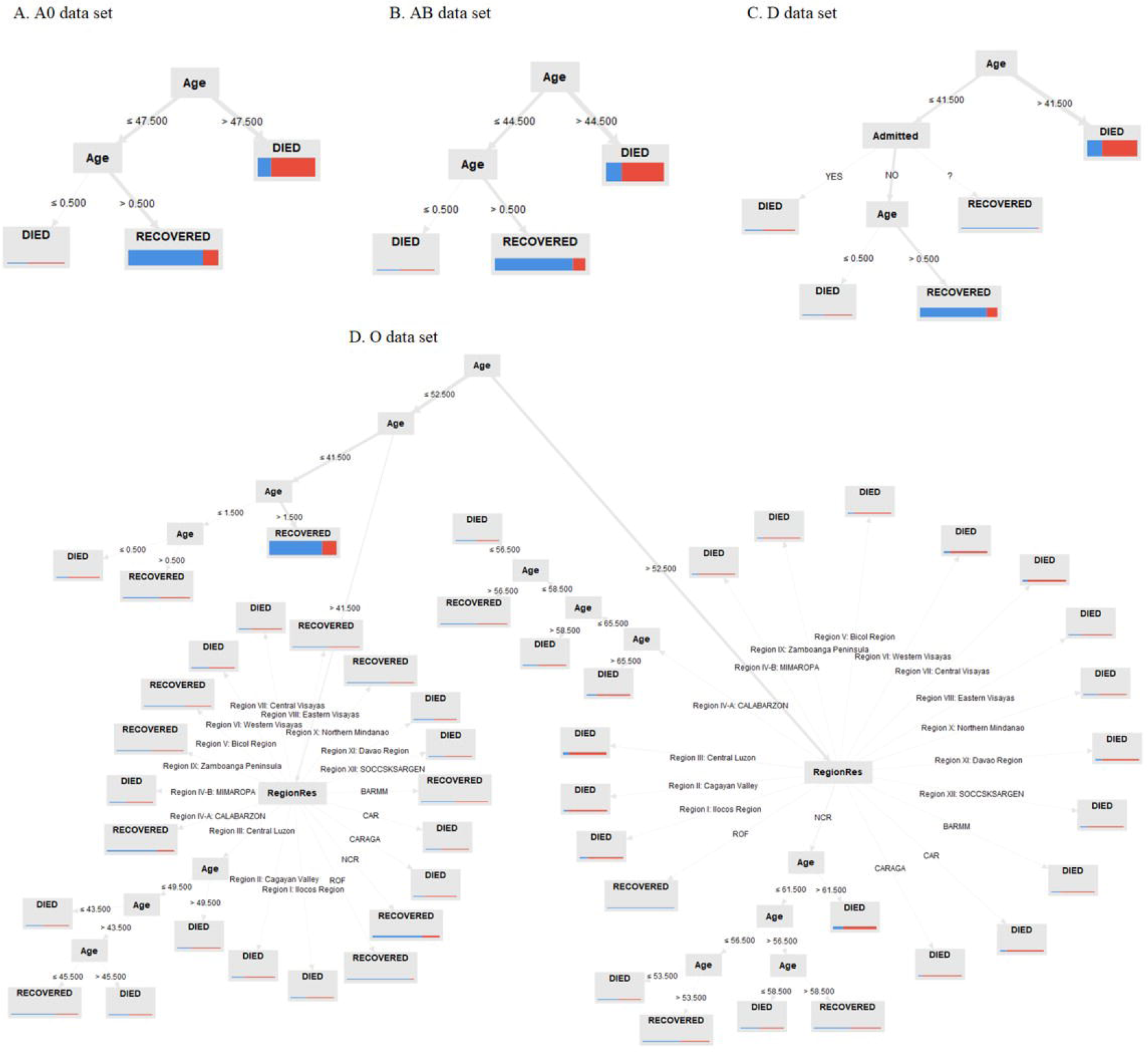
Decision tree for predicted outcomes of reported cases (recovered or died) by data set from the Philippines COVID Data Drop from September 24, 2022^a^ ^a^Relevant attributes identified by the model are shown inside the branches. The predominant outcome per leaf node is identified (either RECOVERED or DIED), with the coloured bars underneath illustrating horizontal stacked bars of the predominant outcome per leaf (RECOVERED=blue, DIED=red). The width of the bars represents the relative number of cases in each leaf as compared with the total cases in the modeling dataset, while the thickness of each arrow illustrates the relative number of cases on each branch as compared with the total cases in the modeling dataset.

The O data set had eight levels, but the number of node splits ranged between two and 16 (**Fig. 3D**). The root node was *Age* with a split criterion of 52.5 years. Cases with *Age* <= 52.5 years were further split according to *Age* <= 41.5 years, with 77.02% of those less than 41.5 years recovering. Cases with *Age* between 41.5 and 52.5 were split into their region of residence, with the majority outcome = DIED for those residing in Regions I, II, III, IV-B, VI, VII, XI, XII, XIII as well as in CAR. Cases with *Age* > 52.5 years were split into region of residence, with majority of cases from any region dying except for repatriate overseas Filipinos. **Supplementary Information B.5** provides the full information of the decision tree models, including the actual number of cases and outcomes per leaf.

## DISCUSSION

We generated four different decision tree models corresponding to the different predominant COVID-19 strain and variants in the Philippines, with age being the root node for all models. The A0 and AB data sets generated simple and similar decision trees with only age as the significant attribute, while the D data set model incorporated admission history as an additional attribute. The O data set generated a more complicated decision tree which incorporated age, admission history and region of residence of the cases into the model. Machine learning models such as decision trees have been used in analyzing trends in COVID-19 data, including in epidemiological modeling^21^ and prediction of disease prognosis.^17,20^

Reported COVID-19 cases in the Philippines reached almost 4 million cases as of September 24, 2022, with most cases occurring during the Delta and Omicron waves despite the relatively shorter duration of these waves compared with the first infection wave from the ancestral strain. SARS-CoV-2 variants have shown increasing transmissibility compared to previous ones, with the Delta and Omicron variants reaching R_0_ of 7 and 10, respectively, compared to 2.5 of the ancestral strain.^8,22^ Other studies reported that the Omicron variant was up to 3.7 times more transmissible compared to the Delta variant and is primarily due to its ability for immune evasion and reinfection regardless of vaccination status and previous infection^23,24^ mainly due to an enhanced viral replication efficiency in the bronchus.^25^

Previous studies have found that the severity among the ancestral, Alpha and Delta variants are comparable,^11,18^ but the severity of the Omicron variant has consistently been lower compared to the other variants.^11,24,26,27^ This may be due to lower replication competence of the Omicron variant in the lung parenchyma.^25^ These findings were consistent with our study: our calculated CFR during the Omicron wave was 65%, 68% and 75% lower than those of the Alpha/Beta, Delta and ancestral waves, respectively. Earlier studies on sex differentials in COVID-19 mortality (i.e., males tend to have higher CFRs)^20,28^ also confirm our results regardless of COVID-19 variant.

In our study, age is the main predictor of our defined outcome for reported COVID-19 cases. Older age groups tend to have higher case fatality rates regardless of predominant COVID-19 variant. This general trend has been documented in previous studies.^1,6,11,29,30^ However, we noticed a pattern similar to a previous Philippine study^20^ on cases of the ancestral variant: the CFRs of the lowest age group (i.e., 0-4 years) tend to be up to 6 times the CFR of the baseline (i.e., 18-29 years), with the lowest CFRs seen in the 5-17 age group. The US Centers for Disease Control and Prevention^29^ shows a generally increasing trend in CFR but a study by Khera et al. (2021) supports our findings and attributed this “U-shaped” phenomenon to several factors such as children having differential expression of ACE-2 receptors, more robust innate immune system (except for newborns), and lesser exposure due to public health measures.^31^

Our decision tree models showed several results. First, among all the attributes included in our models and consistent with our descriptive analysis, age is the most important predictor of mortality. Previous machine learning models on COVID-19 mortality^20,32^ confirm this finding, suggesting that in the absence of clinical data in surveillance data sets, age remains an important factor. Second, the similarities between the A0 and AB models suggest that earlier in the pandemic, the impact of the two waves in the general population may have been similar. During these times, large portions of the population in the country were still under COVID-19 lockdowns and vaccinations had barely started.^33,34^ These events may have limited the population’s exposure to the virus and to COVID-19 vaccines which may suggest that during the early months of the pandemic, internal biological factors such as age-related immunosenescence and presence of comorbidities are bigger factors in prognosis compared to natural or acquired immunity.^1^ Third, the D model incorporated history of admission as a splitting criterion, similar to a previous study.^20^ The previous national guidelines for hospitalization of COVID-19 patients prioritizes admission of only severe and critical COVID-19 cases,^20,35^ and this may have been exacerbated by the sudden influx of COVID-19 cases during the Delta wave as reported in this study.

Fourth, the incorporation of the attribute *RegionRes* (region of residence) in the O data set model is quite novel. A previous study^20^ of the early COVID-19 ancestral wave in the Philippines did not include geopolitical classifications in the model and was consistent with our A0, AB and D models. Our current O model suggests that there may be different impacts of the Omicron variant among different regions in the Philippines. Literature regarding regional differences in COVID-19 CFRs are limited, but previous studies recognized the association of transmission or mortality rates with differences in health care system factors such as number of available hospital beds,^10,36,37^ length and severity of lockdowns, population or industrial composition,^37,38^ and previous infection or vaccination rates.^5,7,12,27^ Repatriated overseas Filipinos, on the other hand, are only allowed to return to the country if they are well enough to travel,^39^ hence the lower CFR among this cohort regardless of infection wave.

Our study has several limitations. Since the data set is publicly available surveillance data, it did not include clinical factors that are associated with COVID-19 mortality such as presence of comorbidities, vaccination status and sociodemographic information. Our categorization of cases according to infection waves was also based on the predominant variant during the date of confirmation of infection and not based on genetic sequencing. Additionally, these dates may have also been delayed. These factors could have led to improper classification of reported cases, particularly those whose reported dates were near the boundaries of our infection wave timelines.

Surveillance data sets during the pandemic are often imbalanced in that the number of recoveries vastly outnumber reported deaths. We used undersampling techniques to control for this imbalance. The models we generated generally had high AUC and sensitivity, with the naïve Bayes and the decision tree models mostly having the highest AUC and sensitivity across the different data sets, respectively. Higher sensitivity is often preferred in inherently imbalanced data sets.^40^ We utilized similar techniques from a previous study^20^ to reduce overfitting: removing irrelevant or highly correlated attributes, enabling pre-pruning and pruning during model training, and optimizing the hyperparameters for the highest sensitivity.

In conclusion, our study highlights the observable changes in COVID-19 transmissibility and case fatality rates depending on the infection timeline and predominant SARS-CoV-2 variant, with the mortality pattern of the Omicron variant being significantly different from the preceding variants. Our findings also reinforce the strong influence of increasing age in predicting COVID-19 outcomes regardless of SARS-CoV-2 variant. The models that we generated highlight the need for up-to-date and stratified policies especially during viral epidemics and pandemics. We recommend future research to apply similar analysis of publicly available surveillance data to monitor emerging or ongoing outbreaks.

## Data Availability

All data used in the present study are available online at https://doh.gov.ph/covid19tracker

## Acknowledgements

The authors would like to thank the San Beda Research and Development Center for the overall support to the study.

## Ethics statement

The study was reviewed and approved by the San Beda University Research Ethics Board on October 21, 2022 under the study protocol code SBU-RED 2022-020. The study adhered to the TRIPOD checklist for prediction model development.

