## Supplementary Information for "COVID-19 infection wave mortality from surveillance data in the Philippines using machine learning"

**Supplementary Information A – Exploratory data pre-processing**

The steps in manual exploratory analysis of the raw dataset are outlined below. Decisions were made by unanimous voting from the authors.

1. The raw dataset contained a total of 3,934,777 cases with 22 attributes. Listed below are the attributes, their operational definitions, and their final disposition after our exploratory analysis:

| # | **Attribute** | **Operational Definition (DOH, 2020)** | **Disposition** |
| --- | --- | --- | --- |
| 1 | CaseCode | Random code assigned for labelling cases; does not equate to the unique case number assigned by DOH | Dropped due to ID-ness |
| 2 | Age* | Age | Included in exploratory dataset |
| 3 | AgeGroup | Five-year age group | Recoded to new attribute *Age_Group* based on US CDC classification using attribute *Age*. Not included in final dataset but analyzed for descriptive statistics. |
| 4 | Sex* | Sex | Included in exploratory dataset |
| 5 | DateSpecimen | Date when specimen was collected | Dropped due to author consensus based on literature review, multiple missing values, ambiguous coding |
| 6 | DateResultRelease | Date of release of result | Dropped due to author consensus based on literature review, multiple missing values, ambiguous coding |
| 7 | DateRepConf** | Date publicly announced as confirmed case | Included in exploratory dataset as reference for timeline of infection variants but not included in final dataset |
| 8 | DateDied | Date died | Dropped due to author consensus based on literature review, multiple missing values, ambiguous coding |
| 9 | DateRecover | Date recovered | Dropped due to author consensus based on literature review, multiple missing values, ambiguous coding |
| 10 | RemovalType* | Type of removal (recovery or death) | Included in final dataset and role was set as “label” or predictor. Only cases without missing values for *RemovalType* were included in the exploratory dataset. |
| 11 | Admitted* | Binary variable indicating patient has been admitted to hospital | Included in exploratory dataset |
| 12 | RegionRes* | Region of residence | Included in exploratory dataset |
| 13 | ProvRes* | Province of residence | Included in exploratory dataset |
| 14 | CityMunRes* | City of residence | Included in exploratory dataset |
| 15 | BarangayRes* | Barangay of residence | Included in exploratory dataset |
| 16 | CityMuniPSGC | Philippine Standard Geographic Code of Municipality or City of Residence | Dropped due to high correlation with *CityMunRes* |
| 17 | BarangayPSGC | Philippine Standard Geographic Code of Barangay of Residence | Dropped due to high correlation with *BarangayRes* |
| 18 | HealthStatus | Known current health status of patient (asymptomatic, mild, severe, critical, died, recovered) | Dropped due to high correlation with *RemovalType* |
| 19 | Quarantined* | Ever been home quarantined, not necessarily currently in home quarantine | Included in exploratory dataset |
| 20 | DateOnset | Date of Onset of Symptoms | Dropped due to author consensus based on literature review, multiple missing values, ambiguous coding |
| 21 | Pregnanttab* | Yes/No if patient is pregnant at any point during COVID-19 condition | Included in exploratory dataset |
| 22 | Validation Status | Text column for comments | Dropped due to free text values and no operational definition |

2. *AgeGroup* was recoded as *Age_Group* to reclassify *Age* into nine bins based on the US CDC classification for descriptive statistics.
3. Missing values for *Pregnanttab* were recoded as “(N/A)” for male cases to distinguish from actual missing values for female cases.
4. Missing values for *BarangayRes*, *CityMunRes* and *ProvRes* were recoded as “ROF” for all cases where *RegionRes*=“ROF.”
5. Cases with missing values for *Age* were dropped from the datasets since literature suggests high importance of this attribute and replacement of missing values may influence the model.
6. The exploratory dataset was then filtered to select only cases with no missing values for *RemovalType* to generate the final dataset. The final dataset (which can be accessed here: <https://osf.io/j478p/?view_only=e5a411cc921a4ac79dd019562d54301d>) consisted of 3,896,206 (99.02%) and 10 attributes including one label attribute (*RemovalType*).
7. The final dataset was then split into four (4) timeline datasets according to COVID-19 surges timelines as reported in the values of *DateRepConf*:
   a. Ancestral strain (A0) dataset: *DateRepConf* < = Feb 28, 2021
   b. Alpha/Beta variants (AB) dataset: Feb 28, 2021 < *DateRepConf* <= June 30, 2021
   c. Delta variant (D) dataset: June 30, 2021 < *DateRepConf* <= Nov 30, 2021
   d. Omicron variant (O) dataset: *DateRepConf* *>* Nov 30, 2021
8. The total number of cases per timeline dataset is listed below:
   a. A0 dataset = 571,905 (RECOVERED=557,657; DIED=14,248; CFR=2.49%)
   b. AB dataset = 835,779 (RECOVERED=821,104; DIED=14,675; CFR=1.76%)
   c. D dataset = 1,414,651 (RECOVERED=1,387,359; DIED=27,292; CFR=1.93%)
   d. O dataset = 1,073,872 (RECOVERED=1,067,374; DIED=6,497; CFR=0.61%)
9. Each timeline datasets (A0, AB, D, O) were individually subjected to the model generation methodology below (Supplementary Information B).

**Supplementary Information B – Model generation**

The steps in machine learning attribute selection, random undersampling and hyperparameter optimizations are outlined below. All processes were conducted using RapidMiner Studio 9.10.008 (rev: 68db53, platform: WIN64).

1. The attribute *RemovalType* was labelled as the outcome to generate the modelling dataset. All random processes were standardized using the random seed “2022”.
2. Attribute selection was done on each training dataset via feature weighting of the nine non-label attributes using the operators *weightbyGiniIndex, weightbyInformationGain* and *weightbyInformationGainRatio*. This was followed by majority voting as the rank aggregation strategy.

Here are the results of the feature weights operators used in feature selection:

Ancestral strain (A0) dataset:

| **Attribute** | **Information Gain** | **Information Gain Ratio** | **Gini Index** |
| --- | --- | --- | --- |
| Sex | <0.001 | <0.001 |  |
| Pregnanttab | <0.001 | <0.001 | <0.001 |
| Quarantined | <0.001 | <0.001 | <0.001 |
| RegionRes | 0.003 | <0.001 | <0.001 |
| ProvRes | 0.003 | <0.001 | <0.001 |
| CityMunRes | 0.006 | <0.001 |  |
| Admitted | 0.013 | 0.038 | 0.002 |
| BarangayRes | 0.017 | 0.002 |  |
| Age | 0.027 | 0.072 | 0.003 |

Alpha + Beta variant (AB) dataset:

| **Attribute** | **Information Gain** | **Information Gain Ratio** | **Gini Index** |
| --- | --- | --- | --- |
| Sex | <0.001 | <0.001 |  |
| Pregnanttab | <0.001 | <0.001 |  |
| Quarantined | <0.001 | <0.001 |  |
| RegionRes | 0.002 | <0.001 | <0.001 |
| ProvRes | 0.003 | <0.001 | <0.001 |
| CityMunRes | 0.005 | <0.001 |  |
| Admitted | 0.006 | 0.026 | 0.001 |
| BarangayRes | 0.017 | 0.002 |  |
| Age | 0.018 | 0.05 | 0.001 |

Delta variant (D) dataset:

| **Attribute** | **Information Gain** | **Information Gain Ratio** | **Gini Index** |
| --- | --- | --- | --- |
| Sex | <0.001 | <0.001 |  |
| Pregnanttab | <0.001 | <0.001 |  |
| Quarantined | <0.001 | 0.001 |  |
| RegionRes | 0.005 | 0.001 | 0.0002 |
| ProvRes | 0.006 | 0.001 | 0.0003 |
| CityMunRes | 0.008 | 0.001 |  |
| Admitted | 0.004 | 0.016 | 0.0004 |
| BarangayRes | 0.018 | 0.002 |  |
| Age | 0.017 | 0.035 | 0.0012 |

Omicron variant (O) dataset:

| **Attribute** | **Information Gain** | **Information Gain Ratio** | **Gini Index** |
| --- | --- | --- | --- |
| Sex | <0.001 | <0.001 |  |
| Pregnanttab | <0.001 | <0.001 |  |
| Quarantined | <0.001 | <0.001 |  |
| RegionRes | 0.002 | <0.001 | 3.16E-05 |
| ProvRes | 0.002 | <0.001 | 4.78E-05 |
| CityMunRes | 0.004 | <0.001 |  |
| Admitted | <0.001 | 0.003 | 1.98E-05 |
| BarangayRes | 0.012 | 0.001 |  |
| Age | 0.006 | 0.038 | 1.55E-04 |

Only *Age* and *Admitted* were considered in the models for datasets A0, AB and D because of relatively high feature weights within operators and consistent rankings across all three operators. For the dataset O, *Age*, *Admitted* and *RegionRes* were included in the model due to relative high feature weights common to information gain ratio and gini index. *ProvRes* was not included in the model for dataset O because *RegionRes* can be inferred from *ProvRes* (i.e., a specific province is located within a defined region) and the inclusion of both attributes may only complicate the model without adding information.

3. The subprocess *optimizeParameters(Grid)* was used to perform grid optimization of the hyperparameters for the decision tree operator *decisionTree* as well as the threshold operator *createThreshold* using the following parameters:

- Decision Tree (XV).criterion: gini_index, gain_ratio
- Decision Tree (XV).minimal_gain: min=0.0, max=0.1, steps=5, scale=linear
- Decision Tree (XV).minimal_leaf_size: min=10, max=110, steps=10, scale=linear
- Decision Tree (XV).minimal_size_for_split: min=10, max=110, steps=10, scale=linear
- Decision Tree (XV).maximal_depth: min=1, max=10, steps=5, scale=linear

The subprocess ran using a cross-validation operator (see step 4 below) to maximize sensitivity. The resulting optimized hyperparameters were as follows, and were used in the final training:

| **Dataset** | **criterion** | **minimal_gain** | **minimal_leaf_size** | **minimal_size_for_split** | **maximal_depth** |
| --- | --- | --- | --- | --- | --- |
| A0 | gain_ratio | 0.08 | 50 | 10 | 4 |
| AB | gain_ratio | 0.08 | 50 | 10 | 4 |
| D | gain_ratio | 0.08 | 50 | 10 | 4 |
| O | gini_index | 0 | 10 | 90 | 10 |

4. The cross-validation operator *crossValidation* was used to train and validate the final datasets using the decision tree model with the optimized *decisionTree* and *createThreshold* hyperparameters generated using the following specifications:
   a. The operator used a five-fold stratified sampling cross-validation with the positive class set as *RemovalType*=DIED.
   b. The main performance metric used was area under the receiver operating characteristic curve (AUC). Other performance metrics were also generated: accuracy, F-score, sensitivity and specificity.
   c. Training dataset: Random undersampling (RUS) was done only on the training dataset for each fold in the cross-validation due to imbalanced values for the label/predictor (*RemovalType*). RUS was done using the *sample* operator to a) select all cases with *RemovalType*=DIED (see below), and b) randomly select cases with *RemovalType*=RECOVERED using simple random sampling to achieve a 1:1 RECOVERED:DIED ratio. This training dataset was used to generate the decision tree models per fold.
   d. Testing dataset: All cases (see below) in the testing dataset were used to validate the model for each fold in the cross-validation. This step ensures that the testing dataset is not influenced by the training dataset RUS, and that the testing dataset represents real-world data.

| **Dataset** | **n for Training per fold** | | **n for Testing per fold** |
| --- | --- | --- | --- |
|  | ***RemovalType*= DIED** | ***RemovalType*= RECOVERED (RUS)** |  |
| A0 | 11,399 | 11,399 | 214,774 |
| AB | 11,740 | 11,740 | 167,156 |
| D | 21,834 | 21,834 | 282,930 |
| O | 5,198 | 5,198 | 114,381 |

**NOTE: n for training used in RUS was calculated by multiplying the N of each dataset by 4/5 while the n for testing was calculated by dividing the N of each dataset by 5 (due to 5-fold cross-validation using 4 subsets as the training dataset and the remaining subset as the testing dataset).*

5. The aggregated decision tree models generated by the cross-validation process using the final datasets were also extracted. The illustrative figures of the decision tree are shown in **Figure 3** while the text description of each node and the models' attribute weights are shown below:
   a. A0 dataset

**Tree:**

```
Age > 47.500: DIED {RECOVERED=2932, DIED=9600}
Age ≤ 47.500
|   Age > 0.500: RECOVERED {RECOVERED=8438, DIED=1743}
|   Age ≤ 0.500: DIED {RECOVERED=29, DIED=56}
```

**Weights:**

| Attribute | Weight |
| --- | --- |
| Age | 1 |

**Performance Vector:**

```
accuracy: 74.06% +/- 0.79% (micro average: 74.06%)
ConfusionMatrix:
True:	RECOVERED	DIED
RECOVERED:	411627	2300
DIED:	146030	11948
classification_error: 25.94% +/- 0.79% (micro average: 25.94%)
ConfusionMatrix:
True:	RECOVERED	DIED
RECOVERED:	411627	2300
DIED:	146030	11948
AUC: 0.789 +/- 0.004 (micro average: 0.789) (positive class: DIED)
precision: 7.57% +/- 0.21% (micro average: 7.56%) (positive class: DIED)
ConfusionMatrix:
True:	RECOVERED	DIED
RECOVERED:	411627	2300
DIED:	146030	11948
recall: 83.86% +/- 0.35% (micro average: 83.86%) (positive class: DIED)
ConfusionMatrix:
True:	RECOVERED	DIED
RECOVERED:	411627	2300
DIED:	146030	11948
f_measure: 13.88% +/- 0.35% (micro average: 13.87%) (positive class: DIED)
ConfusionMatrix:
True:	RECOVERED	DIED
RECOVERED:	411627	2300
DIED:	146030	11948
sensitivity: 83.86% +/- 0.35% (micro average: 83.86%) (positive class: DIED)
ConfusionMatrix:
True:	RECOVERED	DIED
RECOVERED:	411627	2300
DIED:	146030	11948
specificity: 73.81% +/- 0.81% (micro average: 73.81%) (positive class: DIED)
ConfusionMatrix:
True:	RECOVERED	DIED
RECOVERED:	411627	2300
DIED:	146030	11948
```

b. AB dataset

**Tree:**

```
Age > 44.500: DIED {RECOVERED=3877, DIED=10444}
Age ≤ 44.500
|   Age > 0.500: RECOVERED {RECOVERED=7836, DIED=1256}
|   Age ≤ 0.500: DIED {RECOVERED=27, DIED=40}
```

**Weights:**

| Attribute | Weight |
| --- | --- |
| Age | 1 |

**Performance Vector:**

```
accuracy: 68.36% +/- 1.80% (micro average: 68.36%)
ConfusionMatrix:
True:	RECOVERED	DIED
RECOVERED:	558380	1756
DIED:	262724	12919
classification_error: 31.64% +/- 1.80% (micro average: 31.64%)
ConfusionMatrix:
True:	RECOVERED	DIED
RECOVERED:	558380	1756
DIED:	262724	12919
AUC: 0.781 +/- 0.004 (micro average: 0.781) (positive class: DIED)
precision: 4.70% +/- 0.20% (micro average: 4.69%) (positive class: DIED)
ConfusionMatrix:
True:	RECOVERED	DIED
RECOVERED:	558380	1756
DIED:	262724	12919
recall: 88.03% +/- 1.21% (micro average: 88.03%) (positive class: DIED)
ConfusionMatrix:
True:	RECOVERED	DIED
RECOVERED:	558380	1756
DIED:	262724	12919
f_measure: 8.91% +/- 0.35% (micro average: 8.90%) (positive class: DIED)
ConfusionMatrix:
True:	RECOVERED	DIED
RECOVERED:	558380	1756
DIED:	262724	12919
sensitivity: 88.03% +/- 1.21% (micro average: 88.03%) (positive class: DIED)
ConfusionMatrix:
True:	RECOVERED	DIED
RECOVERED:	558380	1756
DIED:	262724	12919
specificity: 68.00% +/- 1.85% (micro average: 68.00%) (positive class: DIED)
ConfusionMatrix:
True:	RECOVERED	DIED
RECOVERED:	558380	1756
DIED:	262724	12919
```

c. D dataset

**Tree:**

```
Age > 41.500: DIED {RECOVERED=8165, DIED=19244}
Age ≤ 41.500
|   Admitted = ?: RECOVERED {RECOVERED=130, DIED=3}
|   Admitted = NO
|   |   Age > 0.500: RECOVERED {RECOVERED=13232, DIED=2084}
|   |   Age ≤ 0.500: DIED {RECOVERED=83, DIED=104}
|   Admitted = YES: DIED {RECOVERED=224, DIED=399}
```

**Weights:**

| Attribute | Weight |
| --- | --- |
| Age | 0.666 |
| Admitted | 0.334 |

**Performance Vector:**

```
accuracy: 66.15% +/- 2.73% (micro average: 66.15%)
ConfusionMatrix:
True:	RECOVERED	DIED
RECOVERED:	911900	3360
DIED:	475459	23932
classification_error: 33.85% +/- 2.73% (micro average: 33.85%)
ConfusionMatrix:
True:	RECOVERED	DIED
RECOVERED:	911900	3360
DIED:	475459	23932
AUC: 0.769 +/- 0.006 (micro average: 0.769) (positive class: DIED)
precision: 4.81% +/- 0.27% (micro average: 4.79%) (positive class: DIED)
ConfusionMatrix:
True:	RECOVERED	DIED
RECOVERED:	911900	3360
DIED:	475459	23932
recall: 87.69% +/- 1.77% (micro average: 87.69%) (positive class: DIED)
ConfusionMatrix:
True:	RECOVERED	DIED
RECOVERED:	911900	3360
DIED:	475459	23932
f_measure: 9.12% +/- 0.48% (micro average: 9.09%) (positive class: DIED)
ConfusionMatrix:
True:	RECOVERED	DIED
RECOVERED:	911900	3360
DIED:	475459	23932
sensitivity: 87.69% +/- 1.77% (micro average: 87.69%) (positive class: DIED)
ConfusionMatrix:
True:	RECOVERED	DIED
RECOVERED:	911900	3360
DIED:	475459	23932
specificity: 65.73% +/- 2.82% (micro average: 65.73%) (positive class: DIED)
ConfusionMatrix:
True:	RECOVERED	DIED
RECOVERED:	911900	3360
DIED:	475459	23932
```

d. O dataset

**Tree:**

```
Age > 52.500
|   RegionRes = BARMM: DIED {RECOVERED=6, DIED=21}
|   RegionRes = CAR: DIED {RECOVERED=31, DIED=187}
|   RegionRes = CARAGA: DIED {RECOVERED=9, DIED=100}
|   RegionRes = NCR
|   |   Age > 61.500: DIED {RECOVERED=151, DIED=512}
|   |   Age ≤ 61.500
|   |   |   Age > 56.500
|   |   |   |   Age > 58.500: RECOVERED {RECOVERED=57, DIED=49}
|   |   |   |   Age ≤ 58.500: DIED {RECOVERED=39, DIED=50}
|   |   |   Age ≤ 56.500
|   |   |   |   Age > 53.500: RECOVERED {RECOVERED=60, DIED=45}
|   |   |   |   Age ≤ 53.500: DIED {RECOVERED=21, DIED=24}
|   RegionRes = ROF: RECOVERED {RECOVERED=10, DIED=0}
|   RegionRes = Region I: Ilocos Region: DIED {RECOVERED=47, DIED=210}
|   RegionRes = Region II: Cagayan Valley: DIED {RECOVERED=28, DIED=258}
|   RegionRes = Region III: Central Luzon: DIED {RECOVERED=88, DIED=527}
|   RegionRes = Region IV-A: CALABARZON
|   |   Age > 65.500: DIED {RECOVERED=50, DIED=179}
|   |   Age ≤ 65.500
|   |   |   Age > 58.500: DIED {RECOVERED=35, DIED=65}
|   |   |   Age ≤ 58.500
|   |   |   |   Age > 56.500: RECOVERED {RECOVERED=21, DIED=17}
|   |   |   |   Age ≤ 56.500: DIED {RECOVERED=32, DIED=36}
|   RegionRes = Region IV-B: MIMAROPA: DIED {RECOVERED=12, DIED=62}
|   RegionRes = Region IX: Zamboanga Peninsula: DIED {RECOVERED=7, DIED=46}
|   RegionRes = Region V: Bicol Region: DIED {RECOVERED=13, DIED=75}
|   RegionRes = Region VI: Western Visayas: DIED {RECOVERED=57, DIED=333}
|   RegionRes = Region VII: Central Visayas: DIED {RECOVERED=45, DIED=350}
|   RegionRes = Region VIII: Eastern Visayas: DIED {RECOVERED=17, DIED=41}
|   RegionRes = Region X: Northern Mindanao: DIED {RECOVERED=25, DIED=47}
|   RegionRes = Region XI: Davao Region: DIED {RECOVERED=41, DIED=228}
|   RegionRes = Region XII: SOCCSKSARGEN: DIED {RECOVERED=16, DIED=70}
Age ≤ 52.500
|   Age > 41.500
|   |   RegionRes = BARMM: RECOVERED {RECOVERED=6, DIED=6}
|   |   RegionRes = CAR: DIED {RECOVERED=18, DIED=32}
|   |   RegionRes = CARAGA: DIED {RECOVERED=8, DIED=23}
|   |   RegionRes = NCR: RECOVERED {RECOVERED=289, DIED=100}
|   |   RegionRes = ROF: RECOVERED {RECOVERED=15, DIED=1}
|   |   RegionRes = Region I: Ilocos Region: DIED {RECOVERED=21, DIED=37}
|   |   RegionRes = Region II: Cagayan Valley: DIED {RECOVERED=17, DIED=43}
|   |   RegionRes = Region III: Central Luzon
|   |   |   Age > 49.500: DIED {RECOVERED=16, DIED=34}
|   |   |   Age ≤ 49.500
|   |   |   |   Age > 43.500
|   |   |   |   |   Age > 45.500: DIED {RECOVERED=26, DIED=30}
|   |   |   |   |   Age ≤ 45.500: RECOVERED {RECOVERED=26, DIED=13}
|   |   |   |   Age ≤ 43.500: DIED {RECOVERED=14, DIED=21}
|   |   RegionRes = Region IV-A: CALABARZON: RECOVERED {RECOVERED=160, DIED=55}
|   |   RegionRes = Region IV-B: MIMAROPA: DIED {RECOVERED=10, DIED=16}
|   |   RegionRes = Region IX: Zamboanga Peninsula: RECOVERED {RECOVERED=12, DIED=11}
|   |   RegionRes = Region V: Bicol Region: RECOVERED {RECOVERED=13, DIED=8}
|   |   RegionRes = Region VI: Western Visayas: DIED {RECOVERED=35, DIED=56}
|   |   RegionRes = Region VII: Central Visayas: DIED {RECOVERED=22, DIED=58}
|   |   RegionRes = Region VIII: Eastern Visayas: RECOVERED {RECOVERED=9, DIED=8}
|   |   RegionRes = Region X: Northern Mindanao: RECOVERED {RECOVERED=20, DIED=12}
|   |   RegionRes = Region XI: Davao Region: DIED {RECOVERED=38, DIED=41}
|   |   RegionRes = Region XII: SOCCSKSARGEN: DIED {RECOVERED=10, DIED=17}
|   Age ≤ 41.500
|   |   Age > 1.500: RECOVERED {RECOVERED=3408, DIED=917}
|   |   Age ≤ 1.500
|   |   |   Age > 0.500: RECOVERED {RECOVERED=59, DIED=50}
|   |   |   Age ≤ 0.500: DIED {RECOVERED=28, DIED=77}
```

**Weights:**

| Attribute | Weight |
| --- | --- |
| Age | 0.866 |
| Admitted | 0.089 |
| RegionRes | 0.045 |

**Performance Vector:**

```
accuracy: 77.27% +/- 2.34% (micro average: 77.27%)
ConfusionMatrix:
True:	RECOVERED	DIED
RECOVERED:	824859	1532
DIED:	242515	4965
classification_error: 22.73% +/- 2.34% (micro average: 22.73%)
ConfusionMatrix:
True:	RECOVERED	DIED
RECOVERED:	824859	1532
DIED:	242515	4965
AUC: 0.814 +/- 0.014 (micro average: 0.814) (positive class: DIED)
precision: 2.02% +/- 0.14% (micro average: 2.01%) (positive class: DIED)
ConfusionMatrix:
True:	RECOVERED	DIED
RECOVERED:	824859	1532
DIED:	242515	4965
recall: 76.42% +/- 2.82% (micro average: 76.42%) (positive class: DIED)
ConfusionMatrix:
True:	RECOVERED	DIED
RECOVERED:	824859	1532
DIED:	242515	4965
f_measure: 3.93% +/- 0.26% (micro average: 3.91%) (positive class: DIED)
ConfusionMatrix:
True:	RECOVERED	DIED
RECOVERED:	824859	1532
DIED:	242515	4965
sensitivity: 76.42% +/- 2.82% (micro average: 76.42%) (positive class: DIED)
ConfusionMatrix:
True:	RECOVERED	DIED
RECOVERED:	824859	1532
DIED:	242515	4965
specificity: 77.28% +/- 2.37% (micro average: 77.28%) (positive class: DIED)
ConfusionMatrix:
True:	RECOVERED	DIED
RECOVERED:	824859	1532
DIED:	242515	4965
```

6. Similar cross-validation operators were used to generate naïve Bayes and random forest models of all datasets for comparison using similar parameters above. The confusion matrices and other available relevant outputs for both models are shown below and are summarized in **Table 2**:
   a. A0 dataset

Naïve Bayes

**Performance Vector:**

```
accuracy: 80.25% +/- 0.60% (micro average: 80.25%)
ConfusionMatrix:
True:	RECOVERED	DIED
RECOVERED:	447401	2684
DIED:	110256	11564
classification_error: 19.75% +/- 0.60% (micro average: 19.75%)
ConfusionMatrix:
True:	RECOVERED	DIED
RECOVERED:	447401	2684
DIED:	110256	11564
AUC: 0.877 +/- 0.004 (micro average: 0.877) (positive class: DIED)
precision: 9.50% +/- 0.23% (micro average: 9.49%) (positive class: DIED)
ConfusionMatrix:
True:	RECOVERED	DIED
RECOVERED:	447401	2684
DIED:	110256	11564
recall: 81.16% +/- 0.55% (micro average: 81.16%) (positive class: DIED)
ConfusionMatrix:
True:	RECOVERED	DIED
RECOVERED:	447401	2684
DIED:	110256	11564
f_measure: 17.00% +/- 0.36% (micro average: 17.00%) (positive class: DIED)
ConfusionMatrix:
True:	RECOVERED	DIED
RECOVERED:	447401	2684
DIED:	110256	11564
sensitivity: 81.16% +/- 0.55% (micro average: 81.16%) (positive class: DIED)
ConfusionMatrix:
True:	RECOVERED	DIED
RECOVERED:	447401	2684
DIED:	110256	11564
specificity: 80.23% +/- 0.62% (micro average: 80.23%) (positive class: DIED)
ConfusionMatrix:
True:	RECOVERED	DIED
RECOVERED:	447401	2684
DIED:	110256	11564
```

Random Forest

**Weights:**

| Attribute | Weight |
| --- | --- |
| Age | 0.972 |
| Admitted | 0.028 |

**Performance Vector:**

```
accuracy: 74.66% +/- 0.62% (micro average: 74.66%)
ConfusionMatrix:
True:	RECOVERED	DIED
RECOVERED:	415072	2361
DIED:	142585	11887
classification_error: 25.34% +/- 0.62% (micro average: 25.34%)
ConfusionMatrix:
True:	RECOVERED	DIED
RECOVERED:	415072	2361
DIED:	142585	11887
AUC: 0.824 +/- 0.018 (micro average: 0.824) (positive class: DIED)
precision: 7.70% +/- 0.17% (micro average: 7.70%) (positive class: DIED)
ConfusionMatrix:
True:	RECOVERED	DIED
RECOVERED:	415072	2361
DIED:	142585	11887
recall: 83.43% +/- 0.54% (micro average: 83.43%) (positive class: DIED)
ConfusionMatrix:
True:	RECOVERED	DIED
RECOVERED:	415072	2361
DIED:	142585	11887
f_measure: 14.10% +/- 0.28% (micro average: 14.09%) (positive class: DIED)
ConfusionMatrix:
True:	RECOVERED	DIED
RECOVERED:	415072	2361
DIED:	142585	11887
sensitivity: 83.43% +/- 0.54% (micro average: 83.43%) (positive class: DIED)
ConfusionMatrix:
True:	RECOVERED	DIED
RECOVERED:	415072	2361
DIED:	142585	11887
specificity: 74.43% +/- 0.65% (micro average: 74.43%) (positive class: DIED)
ConfusionMatrix:
True:	RECOVERED	DIED
RECOVERED:	415072	2361
DIED:	142585	11887
```

b. AB dataset

Naïve Bayes

**Performance Vector:**

```
accuracy: 77.07% +/- 0.08% (micro average: 77.07%)
ConfusionMatrix:
True:	RECOVERED	DIED
RECOVERED:	632006	2567
DIED:	189098	12108
classification_error: 22.93% +/- 0.08% (micro average: 22.93%)
ConfusionMatrix:
True:	RECOVERED	DIED
RECOVERED:	632006	2567
DIED:	189098	12108
AUC: 0.869 +/- 0.004 (micro average: 0.869) (positive class: DIED)
precision: 6.02% +/- 0.06% (micro average: 6.02%) (positive class: DIED)
ConfusionMatrix:
True:	RECOVERED	DIED
RECOVERED:	632006	2567
DIED:	189098	12108
recall: 82.51% +/- 0.62% (micro average: 82.51%) (positive class: DIED)
ConfusionMatrix:
True:	RECOVERED	DIED
RECOVERED:	632006	2567
DIED:	189098	12108
f_measure: 11.22% +/- 0.11% (micro average: 11.22%) (positive class: DIED)
ConfusionMatrix:
True:	RECOVERED	DIED
RECOVERED:	632006	2567
DIED:	189098	12108
sensitivity: 82.51% +/- 0.62% (micro average: 82.51%) (positive class: DIED)
ConfusionMatrix:
True:	RECOVERED	DIED
RECOVERED:	632006	2567
DIED:	189098	12108
specificity: 76.97% +/- 0.07% (micro average: 76.97%) (positive class: DIED)
ConfusionMatrix:
True:	RECOVERED	DIED
RECOVERED:	632006	2567
DIED:	189098	12108
```

Random Forest

**Weights:**

| Attribute | Weight |
| --- | --- |
| Age | 0.880 |
| Admitted | 0.120 |

**Performance Vector:**

```
accuracy: 68.76% +/- 1.11% (micro average: 68.76%)
ConfusionMatrix:
True:	RECOVERED	DIED
RECOVERED:	561769	1792
DIED:	259335	12883
classification_error: 31.24% +/- 1.11% (micro average: 31.24%)
ConfusionMatrix:
True:	RECOVERED	DIED
RECOVERED:	561769	1792
DIED:	259335	12883
AUC: 0.798 +/- 0.003 (micro average: 0.798) (positive class: DIED)
precision: 4.74% +/- 0.12% (micro average: 4.73%) (positive class: DIED)
ConfusionMatrix:
True:	RECOVERED	DIED
RECOVERED:	561769	1792
DIED:	259335	12883
recall: 87.79% +/- 0.90% (micro average: 87.79%) (positive class: DIED)
ConfusionMatrix:
True:	RECOVERED	DIED
RECOVERED:	561769	1792
DIED:	259335	12883
f_measure: 8.99% +/- 0.22% (micro average: 8.98%) (positive class: DIED)
ConfusionMatrix:
True:	RECOVERED	DIED
RECOVERED:	561769	1792
DIED:	259335	12883
sensitivity: 87.79% +/- 0.90% (micro average: 87.79%) (positive class: DIED)
ConfusionMatrix:
True:	RECOVERED	DIED
RECOVERED:	561769	1792
DIED:	259335	12883
specificity: 68.42% +/- 1.14% (micro average: 68.42%) (positive class: DIED)
ConfusionMatrix:
True:	RECOVERED	DIED
RECOVERED:	561769	1792
DIED:	259335	12883
```

c. D dataset

Naïve Bayes

**Performance Vector:**

```
accuracy: 74.74% +/- 0.05% (micro average: 74.74%)
ConfusionMatrix:
True:	RECOVERED	DIED
RECOVERED:	1035235	5260
DIED:	352124	22032
classification_error: 25.26% +/- 0.05% (micro average: 25.26%)
ConfusionMatrix:
True:	RECOVERED	DIED
RECOVERED:	1035235	5260
DIED:	352124	22032
AUC: 0.844 +/- 0.003 (micro average: 0.844) (positive class: DIED)
precision: 5.89% +/- 0.04% (micro average: 5.89%) (positive class: DIED)
ConfusionMatrix:
True:	RECOVERED	DIED
RECOVERED:	1035235	5260
DIED:	352124	22032
recall: 80.73% +/- 0.58% (micro average: 80.73%) (positive class: DIED)
ConfusionMatrix:
True:	RECOVERED	DIED
RECOVERED:	1035235	5260
DIED:	352124	22032
f_measure: 10.98% +/- 0.08% (micro average: 10.98%) (positive class: DIED)
ConfusionMatrix:
True:	RECOVERED	DIED
RECOVERED:	1035235	5260
DIED:	352124	22032
sensitivity: 80.73% +/- 0.58% (micro average: 80.73%) (positive class: DIED)
ConfusionMatrix:
True:	RECOVERED	DIED
RECOVERED:	1035235	5260
DIED:	352124	22032
specificity: 74.62% +/- 0.05% (micro average: 74.62%) (positive class: DIED)
ConfusionMatrix:
True:	RECOVERED	DIED
RECOVERED:	1035235	5260
DIED:	352124	22032
```

Random Forest

**Weights:**

| Attribute | Weight |
| --- | --- |
| Age | 0.708 |
| Admitted | 0.292 |

**Performance Vector:**

```
accuracy: 65.32% +/- 2.62% (micro average: 65.32%)
ConfusionMatrix:
True:	RECOVERED	DIED
RECOVERED:	899956	3227
DIED:	487403	24065
classification_error: 34.68% +/- 2.62% (micro average: 34.68%)
ConfusionMatrix:
True:	RECOVERED	DIED
RECOVERED:	899956	3227
DIED:	487403	24065
AUC: 0.779 +/- 0.005 (micro average: 0.779) (positive class: DIED)
precision: 4.72% +/- 0.26% (micro average: 4.71%) (positive class: DIED)
ConfusionMatrix:
True:	RECOVERED	DIED
RECOVERED:	899956	3227
DIED:	487403	24065
recall: 88.18% +/- 1.73% (micro average: 88.18%) (positive class: DIED)
ConfusionMatrix:
True:	RECOVERED	DIED
RECOVERED:	899956	3227
DIED:	487403	24065
f_measure: 8.96% +/- 0.46% (micro average: 8.93%) (positive class: DIED)
ConfusionMatrix:
True:	RECOVERED	DIED
RECOVERED:	899956	3227
DIED:	487403	24065
sensitivity: 88.18% +/- 1.73% (micro average: 88.18%) (positive class: DIED)
ConfusionMatrix:
True:	RECOVERED	DIED
RECOVERED:	899956	3227
DIED:	487403	24065
specificity: 64.87% +/- 2.70% (micro average: 64.87%) (positive class: DIED)
ConfusionMatrix:
True:	RECOVERED	DIED
RECOVERED:	899956	3227
DIED:	487403	24065
```

d. O dataset

Naïve Bayes

**Performance Vector:**

```
accuracy: 80.30% +/- 0.24% (micro average: 80.30%)
ConfusionMatrix:
True:	RECOVERED	DIED
RECOVERED:	857426	1655
DIED:	209948	4842
classification_error: 19.70% +/- 0.24% (micro average: 19.70%)
ConfusionMatrix:
True:	RECOVERED	DIED
RECOVERED:	857426	1655
DIED:	209948	4842
AUC: 0.843 +/- 0.006 (micro average: 0.843) (positive class: DIED)
precision: 2.25% +/- 0.04% (micro average: 2.25%) (positive class: DIED)
ConfusionMatrix:
True:	RECOVERED	DIED
RECOVERED:	857426	1655
DIED:	209948	4842
recall: 74.53% +/- 1.55% (micro average: 74.53%) (positive class: DIED)
ConfusionMatrix:
True:	RECOVERED	DIED
RECOVERED:	857426	1655
DIED:	209948	4842
f_measure: 4.38% +/- 0.08% (micro average: 4.38%) (positive class: DIED)
ConfusionMatrix:
True:	RECOVERED	DIED
RECOVERED:	857426	1655
DIED:	209948	4842
sensitivity: 74.53% +/- 1.55% (micro average: 74.53%) (positive class: DIED)
ConfusionMatrix:
True:	RECOVERED	DIED
RECOVERED:	857426	1655
DIED:	209948	4842
specificity: 80.33% +/- 0.24% (micro average: 80.33%) (positive class: DIED)
ConfusionMatrix:
True:	RECOVERED	DIED
RECOVERED:	857426	1655
DIED:	209948	4842
```

Random Forest

**Weights:**

| Attribute | Weight |
| --- | --- |
| Age | 0.861 |
| Admitted | 0.084 |
| RegionRes | 0.056 |

**Performance Vector:**

```
accuracy: 78.32% +/- 1.02% (micro average: 78.32%)
ConfusionMatrix:
True:	RECOVERED	DIED
RECOVERED:	836083	1543
DIED:	231291	4954
classification_error: 21.68% +/- 1.02% (micro average: 21.68%)
ConfusionMatrix:
True:	RECOVERED	DIED
RECOVERED:	836083	1543
DIED:	231291	4954
AUC: 0.844 +/- 0.006 (micro average: 0.844) (positive class: DIED)
precision: 2.10% +/- 0.07% (micro average: 2.10%) (positive class: DIED)
ConfusionMatrix:
True:	RECOVERED	DIED
RECOVERED:	836083	1543
DIED:	231291	4954
recall: 76.25% +/- 1.61% (micro average: 76.25%) (positive class: DIED)
ConfusionMatrix:
True:	RECOVERED	DIED
RECOVERED:	836083	1543
DIED:	231291	4954
f_measure: 4.09% +/- 0.13% (micro average: 4.08%) (positive class: DIED)
ConfusionMatrix:
True:	RECOVERED	DIED
RECOVERED:	836083	1543
DIED:	231291	4954
sensitivity: 76.25% +/- 1.61% (micro average: 76.25%) (positive class: DIED)
ConfusionMatrix:
True:	RECOVERED	DIED
RECOVERED:	836083	1543
DIED:	231291	4954
specificity: 78.33% +/- 1.04% (micro average: 78.33%) (positive class: DIED)
ConfusionMatrix:
True:	RECOVERED	DIED
RECOVERED:	836083	1543
DIED:	231291	4954
```

7. The receiver operating characteristics (ROC) curves of the three models were plotted into one graph and is shown in **Figure 2**.

**Supplementary Information C - TRIPOD Checklist: Prediction Model Development**

| **Section/Topic** | **Item** | **Checklist Item** | **Page** |
| --- | --- | --- | --- |
| **Title and abstract** | | | |
| Title | 1 | Identify the study as developing and/or validating a multivariable prediction model, the target population, and the outcome to be predicted. | 1 |
| Abstract | 2 | Provide a summary of objectives, study design, setting, participants, sample size, predictors, outcome, statistical analysis, results, and conclusions. | 1 |
| **Introduction** | | | |
| Background and objectives | 3a | Explain the medical context (including whether diagnostic or prognostic) and rationale for developing or validating the multivariable prediction model, including references to existing models. | 2-3 |
|  | 3b | Specify the objectives, including whether the study describes the development or validation of the model or both. | 2-3 |
| **Methods** | | | |
| Source of data | 4a | Describe the study design or source of data (e.g., randomized trial, cohort, or registry data), separately for the development and validation data sets, if applicable. | 3-4 |
|  | 4b | Specify the key study dates, including start of accrual; end of accrual; and, if applicable, end of follow-up. | 3 |
| Participants | 5a | Specify key elements of the study setting (e.g., primary care, secondary care, general population) including number and location of centres. | 3 |
|  | 5b | Describe eligibility criteria for participants. | 3 |
|  | 5c | Give details of treatments received, if relevant. | n/a |
| Outcome | 6a | Clearly define the outcome that is predicted by the prediction model, including how and when assessed. | 3-4 |
|  | 6b | Report any actions to blind assessment of the outcome to be predicted. | n/a |
| Predictors | 7a | Clearly define all predictors used in developing or validating the multivariable prediction model, including how and when they were measured. | 3-4 |
|  | 7b | Report any actions to blind assessment of predictors for the outcome and other predictors. | n/a |
| Sample size | 8 | Explain how the study size was arrived at. | 3 |
| Missing data | 9 | Describe how missing data were handled (e.g., complete-case analysis, single imputation, multiple imputation) with details of any imputation method. | 3, S2 |
| Statistical analysis methods | 10a | Describe how predictors were handled in the analyses. | 3-4, S3-5 |
|  | 10b | Specify type of model, all model-building procedures (including any predictor selection), and method for internal validation. | 3-4, S3-5 |
|  | 10d | Specify all measures used to assess model performance and, if relevant, to compare multiple models. | 3-4, S4 |
| Risk groups | 11 | Provide details on how risk groups were created, if done. | 3 |
| **Results** | | | |
| Participants | 13a | Describe the flow of participants through the study, including the number of participants with and without the outcome and, if applicable, a summary of the follow-up time. A diagram may be helpful. | 4 |
|  | 13b | Describe the characteristics of the participants (basic demographics, clinical features, available predictors), including the number of participants with missing data for predictors and outcome. | 4 |
| Model development | 14a | Specify the number of participants and outcome events in each analysis. | 4 |
|  | 14b | If done, report the unadjusted association between each candidate predictor and outcome. | n/a |
| Model specification | 15a | Present the full prediction model to allow predictions for individuals (i.e., all regression coefficients, and model intercept or baseline survival at a given time point). | 4-5, S5-15 |
|  | 15b | Explain how to the use the prediction model. | 4-5 |
| Model performance | 16 | Report performance measures (with CIs) for the prediction model. | 14-15 |
| **Discussion** | | | |
| Limitations | 18 | Discuss any limitations of the study (such as nonrepresentative sample, few events per predictor, missing data). | 6 |
| Interpretation | 19b | Give an overall interpretation of the results, considering objectives, limitations, and results from similar studies, and other relevant evidence. | 5-6 |
| Implications | 20 | Discuss the potential clinical use of the model and implications for future research. | 6 |
| **Other information** | | | |
| Supplementary information | 21 | Provide information about the availability of supplementary resources, such as study protocol, Web calculator, and data sets. | 3-5, S2 |
| Funding | 22 | Give the source of funding and the role of the funders for the present study. | Covid Variants_OTHERS.docx |

**Supplementary Information D – RapidMiner processes**

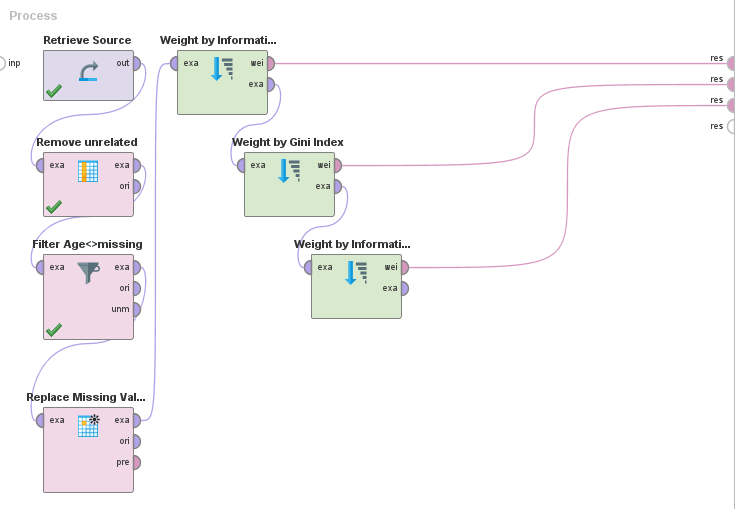

**Figure D.1.** Feature Selection process

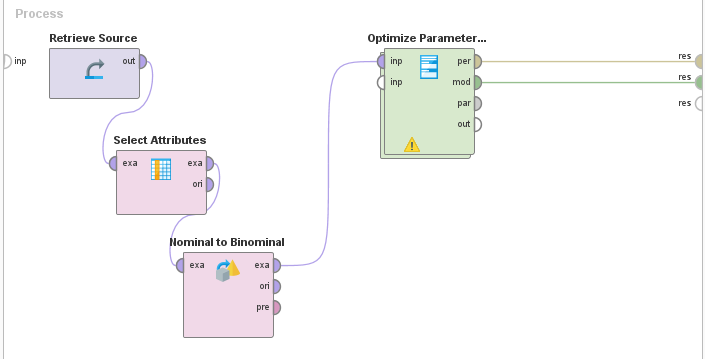

**Figure D.2.** Optimization subprocess

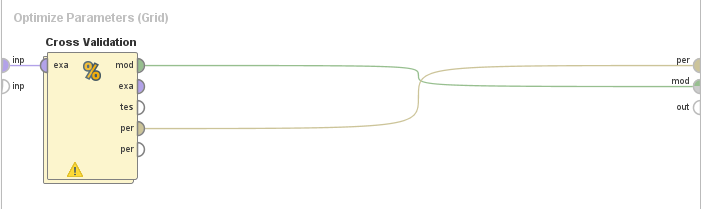

**Figure D.3.** Cross-validation operator

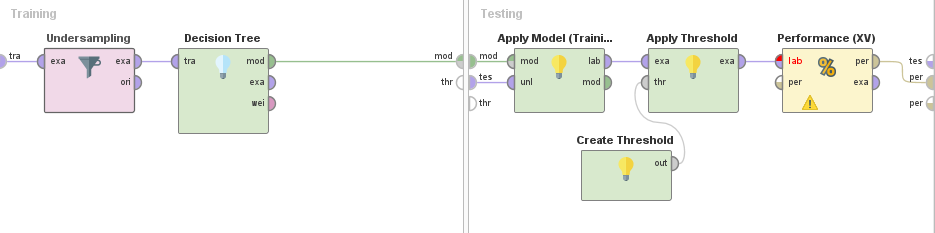

**Figure D.4.** Random undersampling of training sets and model training (left); model testing, application of threshold and performance measurements (right)
